# Neonatal outcomes during the COVID-19 pandemic in New York City

**DOI:** 10.1101/2020.12.20.20248583

**Authors:** Felix Richter, Arielle S. Strasser, Mayte Suarez-Farinas, Shan Zhao, Girish N. Nadkarni, Ethylin Wang Jabs, Katherine Guttmann, Benjamin S. Glicksberg

**Author notes:** **Correspondence:** Benjamin S. Glicksberg, PhD, Icahn School of Medicine at Mount Sinai 770 Lexington Ave., 14^th^Floor, New York, New York 10065, E. Contributed equally (Felix Richter, Arielle S. Strasser). These authors jointly directed this project (Ethylin Wang Jabs, Katherine Guttmann, Benjamin S. Glicksberg).

## Abstract

We explored rates of premature births and neonatal intensive care unit (NICU) admissions at the Mount Sinai Hospital after the implementation of COVID-19 lockdown measures (March 16, 2020) and phase one reopening (June 8, 2020), comparing them to those of the same time periods from 2012-2019. Mount Sinai Hospital is in New York City (NYC), an early epicenter of COVID-19 in the United States, which was heavily impacted by the pandemic during the study period. Among 43,963 singleton births, we observed no difference in either outcome after the implementation of lockdown measures when compared to the same trends in prior years (p=0.09-0.35). Of interest, we observed a statistically significant decrease in premature births after NYC phase one reopening compared to those of the same time period in 2012-2019 across all time windows (p=0.0028-0.049), and a statistically significant decrease in NICU admissions over the largest time window (2.75 months) compared to prior years (p=0.0011).

## INTRODUCTION

New York City (NYC) emerged as the epicenter of the COVID-19 pandemic in the United States, prompting lockdown measures including school closings, stay-at-home orders, and a shift to working from home.[1] These public health interventions were initially instituted to curb transmission of the virus, but have provided insights into associations between lockdown measures and health outcomes. Several reports linked COVID-19 lockdown measures to reduced rates of prematurity, providing possible mechanisms for prematurity prevention.[2–6] To date, this link has not been assessed in NYC, one of the hardest hit and most populous, diverse cities in the United States. Furthermore, neonatal intensive care unit (NICU) admissions, which are related but not exclusively modified by prematurity rates, may also be impacted by lockdown measures. Both of these neonatal outcomes may also be influenced by NYC phase one reopening, which was associated with additional policy and behavioral changes. We therefore assessed if the NYC COVID-19 lockdown measures or NYC phase one reopening were associated with changes in prematurity or NICU admission rates in a large and diverse cohort at the Mount Sinai Hospital.

## METHODS

There were 66,363 neonates born at the Mount Sinai Hospital between January 1, 2012 and November 25, 2020. We excluded 22,400 neonates, the majority of whom had a primary residence outside of NYC and may have experienced different lockdown measures and reopening timelines (**supplemental figure 1**). We also excluded neonates from multiple births and those born to COVID-19 positive mothers, which are potential risk factors for prematurity.[2] We defined prematurity as gestational age <37 weeks and late term as >41+6 weeks, excluding late term neonates from our analysis. Our final cohort totaled 43,963 singleton neonates, including 3,348 live births since the implementation of COVID-19 lockdown measures (**supplemental figure 1**). Data were obtained from electronic health records (**supplemental figure 1**) and research was approved by the Mount Sinai Institutional Review Board. We used March 16, 2020 as the first calendar date of lockdown measures because it had the largest drop in mobility and was the date of NYC’s public school system closure (**supplemental figure 2**).[1] We used June 8, 2020 as the date of reopening because it was the official date of the implementation of NYC phase one reopening.

We used a quasi-experimental difference-in-difference (DiD) logistic regression to test for associations between NYC lockdown measures or reopening and our two primary outcomes, prematurity and NICU admission rates (model in **supplemental methods**). We compared one-, two-, and three-month epochs before and after the implementation of the two pandemic response measures in 2020 to the same short time periods in 2012-2019. Comparing the trends of these short time intervals to the trends in prior years limits the influence of confounding variables, such as seasonal prematurity changes. We assessed these three windows (±1, ±2, and ±3 months) before and after each calendar date to test for sensitivity to time interval choice; for the reopening analysis we limited the largest window to the start date of lockdown (*i.e*. 2.75 months before and after reopening). The total births, premature births, and NICU admissions in each of these analyses is enumerated in **supplemental table 2**. We conducted a sensitivity analysis excluding data within 0.1 and 0.2 months (three and six days, respectively) of each implementation date to test for robustness of date choice. To evaluate the impact of excluding neonate-mother pairs with missing gestational age, we conducted a sensitivity analysis using only data from 2017-2020, during which gestational age missingness was <1%. All statistical analyses were performed in R 3.5.1.

## RESULTS

The characteristics of our cohort are shown in **supplemental table 1**. In our 2020 cohort, 38% of neonates were white, 13% were black, and 6.1% were Asian (**supplemental table 1**). We observed increases in prematurity rates and NICU admissions after the implementation of NYC lockdown measures (March 16, 2020), but these increases were not statistically significant when compared to changes over the same time period in 2012-2019 (**figure 1**, *P*_*interaction*_=0.18-0.35, *P*_*interaction*_=0.09-0.15, respectively). After NYC phase one reopening (June 8, 2020), we observed a decrease in premature births that was statistically significant when compared to 2012-2019 (**figure 2**, *P*_*interaction*_=0.0028-0.049). Consistent with this result, we observed a decrease in NICU admissions with reopening (*P*_*interaction*_=0.0011-0.27). These results were robust to excluding data around the implementation date, as well as restriction to births since 2017 (**supplemental figure 3**).

**Figure 1.**
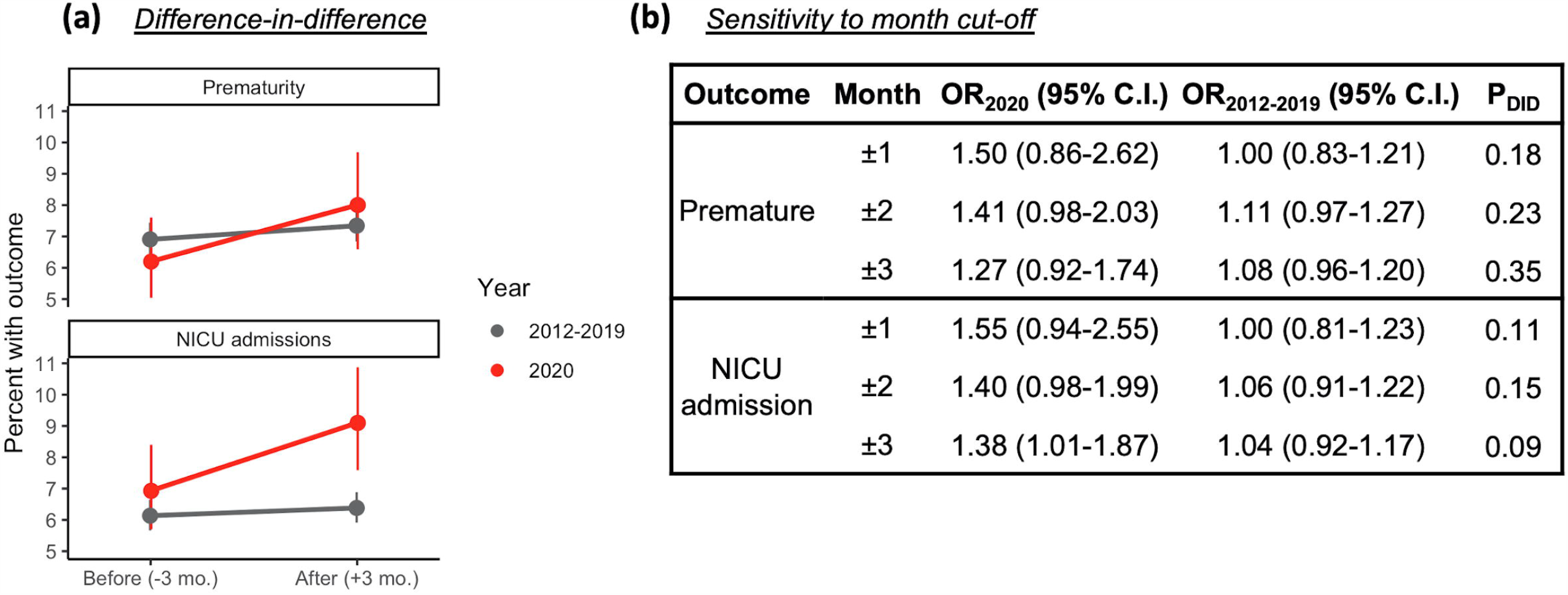
Comparable changes to previous years in prematurity and NICU admission rates at the Mount Sinai Hospital after New York City (NYC) implemented lockdown measures. **(a)** Change in the percent of premature births (GA<37 weeks) (top panel) and NICU admissions (bottom panel) in the three months before and after the implementation of COVID-19 lockdown measures (March 16, 2020), compared to the same time periods in 2012-2019. **(b)** Using a difference-in-difference (DiD) logistic regression, we observed comparable changes of prematurity and NICU admissions after lockdown measure implementation across all month cut-offs (OR = Odds Ratio, 95% CI = 95% Confidence Interval (lower-upper), p-DiD=p-value of the DiD coefficient).

**Figure 2.**
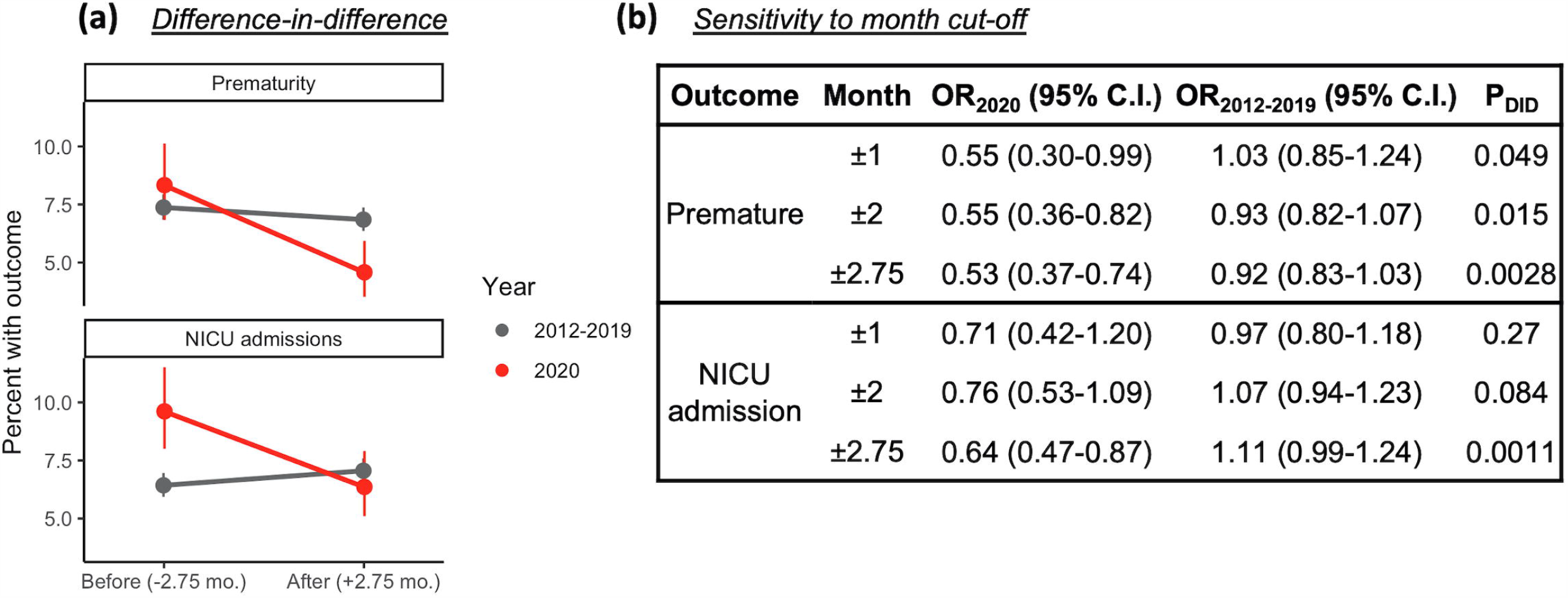
Statistically significant reduction in prematurity and NICU admission rates after the initiation of NYC phase one reopening compared to 2012-2019. **(a)** Change in the percent of premature births (top panel) and NICU admissions (bottom panel) in the 2.75 months before and after NYC phase one reopening (June 8, 2020) compared to the same time periods in 2012-2019 (grey). **(b)** We observed a statistically significant decrease in prematurity and NICU admission rates after reopening when compared to 2012-2019 across all month cut-offs using a difference-in-difference (DiD) logistic regression. We limited our largest window to start at the date of the original lockdown (*i.e*., 2.75 months before reopening). (OR = Odds Ratio, 95% CI = 95% Confidence Interval (lower-upper), p-DiD=p-value of the DiD coefficient).

## DISCUSSION

Studies from the Netherlands, Denmark, Ireland, Italy, and Japan demonstrated a reduction in preterm births following the implementation of lockdown measures.[2–6] In contrast, studies from the United States in California and Philadelphia reported no significant change in preterm births after lockdown.[7,8] In this NYC study, we observed comparable rates in prematurity and NICU admissions after the implementation of lockdown measures when compared to those of the same time period in previous years. Thus far, all reports have observed no change or a reduction in prematurity, and none have shown increases. Our study also assessed prematurity and NICU admission trends after NYC phase one reopening, where we indeed observed a statistically significant reduction in prematurity and NICU admission rates compared to changes after the same calendar date in prior years.

Associations between COVID-19 lockdown and reopening measures with prematurity are multifactorial and affected by patient, health system, sociodemographic, and geographical factors. Our results may reflect changes in obstetric care and shifts in health services. For example, changes in timing of cesarean section or induction in response to maternal or fetal health concerns could impact prematurity rates.[2] Our observations could also be attributable to a delayed impact of NYC lockdown measures on factors previously implicated in prematurity, decreased exposure to non-COVID-19 infections, better air quality, and increased hygienic practices.[2] This delay could be specific to NYC, which experienced an overburdened hospital system and exodus during the earliest days of the pandemic in the United States.[1] Small sample size and absence of information on socioeconomic status, neonatal diagnoses, and maternal risk factors, such as age, parity, and other reproductive and medical conditions, also meant that we were unable to measure the impact of these important determinants on our outcomes of interest. Thus, while we did observe a statistically significant decrease in prematurity and NICU admission rates after reopening, the mechanisms underlying this change are likely multifaceted and warrant further investigation.

## Data Availability

Data are not available due to patient privacy and HIPAA considerations.

## Acknowledgements

Daniel R. Diamond, Shoshana Rosenzweig, Samuel Lee, Nidhi Naik, patients and their families.

