## Supplementary material for "Neonatal outcomes during the COVID-19 pandemic in New York City": Supplement.pdf

**SUPPLEMENTAL METHODS**

We used the following difference-in-difference logistic regression model to test for an association between premature births and the effect of lockdown measures, and separately the subsequent reopening, on non-COVID-affected births:

$$\text{Logit}(p_{premature})=\beta_0+\beta_1*X_{born\_in\_2020}+\beta_2*X_{post\_cut\_off}+\beta_3*X_{born\_in\_2020}*X_{post\_cut\_off}+e_i$$

Here $p_{premature}$ was 1 if the baby was born at gestational age <37 weeks and zero otherwise, born_in_2020 was binary 0/1 if born in the year 2020, post_cut_off was binary 0/1 if born after the date of interest (March 16, 2020 for lockdown or June 8, 2020 for reopening, as separate models). Sensitivity analyses were performed for ±1, ±2, and ±3 month periods before and after lockdown measures and ±1, ±2, and ±2.75 month periods for reopening.

We also assessed the effect of lockdown, and separately the subsequent reopening, on NICU admission rates for non-COVID-affected births with the following model:

$$\text{Logit}(p_{NICU\_admission})=\beta_0+\beta_1*X_{born\_in\_2020}+\beta_2*X_{post\_cut\_off}+\beta_3*X_{born\_in\_2020}*X_{post\_cut\_off}+e_i$$

Here $p_{NICU\_admission}$ was 1 if the baby was admitted to the NICU at birth and zero otherwise, born_in_2020 was binary 0/1 if born in the year 2020 or 2012-2019, post_cut_off was binary 0/1 if born after the date of interest (March 16, 2020 for lockdown or June 8, 2020 for reopening, as separate models).

**SUPPLEMENTARY TABLE**

**Supplemental Table 1. Characteristics of study cohort from 2012-2019 and 2020.**

| | **2012-2019** | **2020** |
|---|---|---|
| Total births | 39,495 | 4468 |
| Term births | 36,736 (93%) | 4,190 (93.78%) |
| Premature births | 2,759 (7.0%) | 278 (6.22%) |
| *Race, neonate* | | |
| American Indian Or Alaska Native | 71 (0.2%) | 37 (0.8%) |
| Asian | 1,417 (3.6%) | 271 (6.1%) |
| Black Or African-American | 4,721 (12%) | 559 (13%) |
| Native Hawaiian Or Pacific Islander | 1,516 (3.8%) | 11 (0.3%) |
| White | 18,960 (48%) | 1,638 (38%) |
| Other | 9,991 (25%) | 1,100 (25%) |
| Unknown | 2,819 (7.1%) | 841 (19%) |

Data are n (%).

**Supplemental Table 2. Total births, premature births, and NICU admissions.**

| Month interval | Year | Pre/post-date | Lockdown measures | | | NYC phase one reopening | | |
|---|---|---|---|---|---|---|---|---|
| | | | Total | Premature | NICU | Total births | Premature | NICU |
| ±1 | 2012-2019 | After | 3049 | 232 | 184 | 3441 | 229 | 216 |
| | 2012-2019 | Before | 3077 | 233 | 186 | 3305 | 215 | 214 |
| | 2020 | After | 401 | 31 | 40 | 396 | 18 | 25 |
| | 2020 | Before | 434 | 23 | 29 | 413 | 33 | 36 |
| ±2 | 2012-2019 | After | 6270 | 485 | 402 | 6918 | 474 | 480 |
| | 2012-2019 | Before | 6202 | 436 | 378 | 6508 | 475 | 424 |
| | 2020 | After | 767 | 68 | 72 | 866 | 40 | 59 |
| | 2020 | Before | 913 | 59 | 63 | 775 | 63 | 68 |
| ±3 or ±2.75* | 2012-2019 | After | 9665 | 710 | 617 | 9525 | 651 | 672 |
| | 2012-2019 | Before | 9184 | 633 | 563 | 8788 | 648 | 565 |
| | 2020 | After | 1187 | 95 | 108 | 1179 | 54 | 75 |
| | 2020 | Before | 1371 | 85 | 95 | 1092 | 91 | 105 |

*±3 months for lockdown measures, ±2.75 months for reopening

**SUPPLEMENTARY FIGURES**

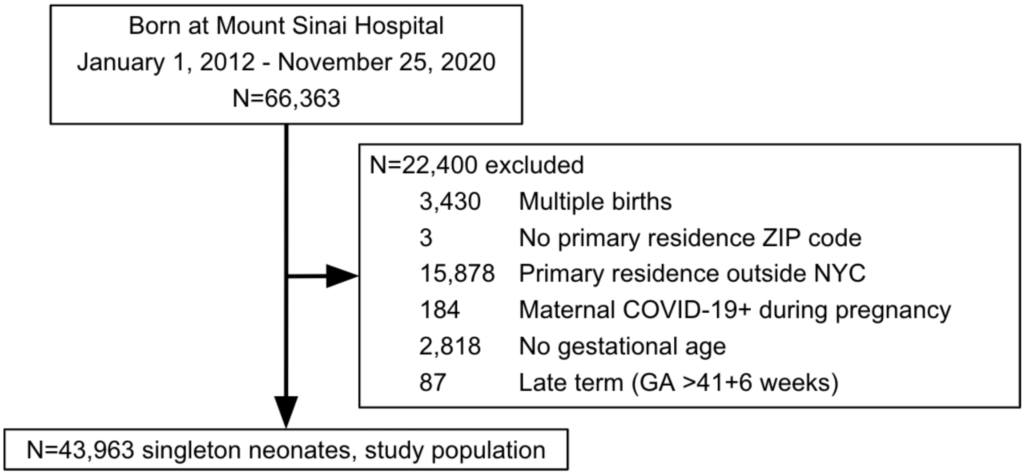

**Supplemental Figure 1.** Consort diagram illustrating patients included in this study.

| | |
|---|---|
| **March 1, 2020** | First reported COVID-19 case in New York State |
| **March 7, 2020** | NY Governor Andrew Cuomo declares a state of emergency |
| | Nursing homes prohibit outside visitors |
| **March 8, 2020** | NYS issues guidelines to avoid densely packed buses, subways, trains |
| **March 10, 2020** | Cuomo orders containment zone in New Rochelle from March 12-25 |
| | National Guard troops deployed to a Health Department post in New Rochelle |
| **March 11, 2020** | WHO declares COVID-19 a global pandemic |
| **March 12, 2020** | Events >500 people must be cancelled or postponed |
| | Broadway closes |
| **March 13, 2020** | President Trump declares a national emergency |
| | Travel ban on non-US citizens travelling from Europe goes into effect |
| **March 15, 2020** | CDC issues guidelines for gatherings <50 people |
| **March 16, 2020** | Closure of NYC public schools |
| | Largest drop in mobility |
| | Physical (“social”) distancing introduced |
| **March 17, 2020** | NYC bars and restaurants close |
| **March 22, 2020** | NYS “PAUSE” program begins |
| | All non-essential workers mandated to stay at home |
| **March 28, 2020** | Non-essential construction halted in NYS |
| **April 15, 2020** | Mandated wear of face masks or coverings in public |
| **April 30, 2020** | NYC subway closures from 1-5 am |
| **May 15, 2020** | Resumption of low risk activities (drive-in-theaters, landscaping, low risk recreational activities) |
| **June 8, 2020** | NYC phase 1 reopening (construction, agriculture, fishing, hunting, curbside retail-pickup, manufacturing, higher education research) |
| **June 22, 2020** | NYC phase 2 reopening (outdoor dining, offices (low capacity), movie theaters, hair salons) |
| **July 6, 2020** | NYC phase 3 reopening (indoor and outdoor dining with six feet distance between tables, personal care businesses, nail salons, spas) |
| **July 19, 2020** | NYC phase 4 reopening (malls, gyms, fitness centers, professional sports with no fans, media production) |
| **September 21, 2020** | Pre-K and special needs classes resume in person learning |
| **September 29, 2020** | Public elementary schools resume in person learning |

**Supplemental Figure 2.** Timeline of pandemic response measures in New York City (NYC). NY = New York, NYS = New York State, WHO = World Health Organization, CDC = Center for Disease Control and Prevention, Pre-K = Pre-kindergarten.

| | | Births ± 0.1 months excluded | | | Births ± 0.2 months excluded | | | Births in 2017-2020 | | |
|---|---|---|---|---|---|---|---|---|---|---|
| **Outcome** | **Month** | **$OR_{2020}$ (95% C.I.)** | **$OR_{2012\text{-}2019}$ (95% C.I.)** | **$P_{DID}$** | **$OR_{2020}$ (95% C.I.)** | **$OR_{2012\text{-}2019}$ (95% C.I.)** | **$P_{DID}$** | **$OR_{2020}$ (95% C.I.)** | **$OR_{2012\text{-}2019}$ (95% C.I.)** | **$P_{DID}$** |
| **Lockdown** | ±1 | 1.42 (0.78-2.57) | 0.93 (0.76-1.14) | 0.19 | 1.37 (0.73-2.58) | 0.87 (0.70-1.08) | 0.19 | 1.501 (0.86-2.62) | 0.92 (0.69-1.24) | 0.13 |
| | ±2 | 1.38 (0.95-2.00) | 1.075 (0.94-1.23) | 0.22 | 1.36 (0.93-2) | 1.052 (0.91-1.21) | 0.22 | 1.41 (0.98-2.03) | 0.98 (0.80-1.21) | 0.093 |
| | ±3 | 1.24 (0.90-1.71) | 1.06 (0.94-1.18) | 0.36 | 1.22 (0.88-1.70) | 1.04 (0.93-1.17) | 0.37 | 1.27 (0.92-1.74) | 1.024 (0.86-1.22) | 0.248 |
| **Reopening** | ±1 | 0.59 (0.32-1.084) | 1.045 (0.86-1.29) | 0.08 | 0.57 (0.29-1.12) | 1.045 (0.84-1.30) | 0.095 | 0.55 (0.30-0.99) | 0.95 (0-.72-1.27) | 0.099 |
| | ±2 | 0.56 (0.37-0.85) | 0.94 (0.82-1.08) | 0.02 | 0.55 (0.36-0.86) | 0.93 (0.81-1.071) | 0.027 | 0.547 (0.36-0.82) | 1.013 (0.82-1.25) | 0.0085 |
| | ±2.75 | 0.54 (0.38-0.77) | 0.93 (0.82-1.04) | 0.004 | 0.53 (0.37-0.76) | 0.92 (0.82-1.03) | 0.005 | 0.53 (0.37-0.75) | 0.94 (0.79-1.13) | 0.0035 |

**Supplemental Figure 3. Sensitivity analyses for lockdown and reopening measures.** Sensitivity to date choice was assessed with difference-in-difference (DiD) logistic regression analyses of prematurity rates excluding births within **(a)** 0.1 months (3 days) and **(b)** 0.2 months (6 days) of lockdown measures or reopening. **(c)** Sensitivity analysis for births from 2017-2020 only, to account for missing gestational age data in 10.2% of babies born in 2012-2016. This analysis only includes neonates born 3 months before March 16, 2017 through November 25, 2020 (<1% of babies missing gestational age). We observed consistent effect sizes and p-values at all month cut-offs. (OR = Odds Ratio, 95% CI = 95% Confidence Interval (lower-upper), p-DiD=p-value of the DiD coefficient).
